# Regression with race-modifiers: towards equity and interpretability

**DOI:** 10.1101/2024.01.04.23300033

**Authors:** Daniel R. Kowal

## Abstract

The pervasive effects of structural racism and racial discrimination are well-established and offer strong evidence that the effects of many important variables on health and life outcomes vary by race. Alarmingly, standard practices for statistical regression analysis introduce racial biases into the estimation and presentation of these race-modified effects. We advocate *abundance-based constraints* (ABCs) to eliminate these racial biases. ABCs offer a remarkable invariance property: estimates and inference for main effects are nearly unchanged by the inclusion of race-modifiers. Thus, quantitative researchers can estimate race-specific effects “for free”—without sacrificing parameter interpretability, equitability, or statistical efficiency. The benefits extend to prominent statistical learning techniques, especially regularization and selection. We leverage these tools to estimate the joint effects of environmental, social, and other factors on 4th end-of-grade readings scores for students in North Carolina (*n* = 27, 638) and identify race-modified effects for racial (residential) isolation, PM_2.5_ exposure, and mother’s age at birth.

---

Health and life outcomes are inextricably linked to race (1, 2). Racial disparities exist in birth outcomes, mortality, disease onset and progression, socioeconomic status, and police-involved deaths, along with many other health and life outcomes (2–4). These disparities persist even after adjusting for socioeconomic status and occur through multiple pathways (1). Structural racism contributes to significant differences in the quality of education, housing, employment opportunities, accumulation of wealth, access to medical care, and treatment in the criminal justice system (1, 2, 5–7). Perceived racial discrimination impacts both mental and physical health through heightened stress responses, health behaviors, and traumatic experiences (8, 9). Thus, rigorous studies of health and life outcomes must carefully consider race as a primary factor.

That race permeates so many aspects of an individual’s life course is a strong indicator that the effects of important factors (*X*) on health and life outcomes (*Y*) may be *race-specific* (10). Regression analysis—the primary statistical tool to quantify how these covariates *X* determine, predict, or associate with an outcome *Y* —must therefore consider *race-modifiers* for *X*. Indeed, there is abundant and growing evidence for race-specific effects, including the effects of red-lining, PM_2.5_ exposure, and cigarette use on mortality risks (11–13); maternal age, poverty, education, and hypertension on infant birthweight, infant mortality, and maternal stroke risk (14–16); education level on multiple health outcomes (17); mood/anxiety disorder on chronic physical health conditions (18); perceived racism on mental health (9); age on allostatic load scores, known as “racial weathering” (19); and the timing of hypertension, insulin resistance, or diabetes onset (20, 21), among many others (22–26). The identification and quantification of race-modified effects are essential to understand and eliminate harmful race disparities in health and life outcomes (27).

To provide context for race-modified effects, we present a regression of 4th end-of-grade reading scores on racial (residential) isolation (RI) and race (Figure 1). The dataset, detailed and reanalyzed subsequently, includes *n* = 27, 638 students in North Carolina (58% non-Hispanic (NH) White, 36% NH Black, 6% Hispanic). RI measures the geographic separation of NH Black individuals and communities from other race groups, and thus is an important measure of structural racism (5, 6, 28–30). The *main-only* (or ANCOVA) model (Figure 1a) includes race only as an additive effect, which restricts the RI effect to be common across race groups. This widely-used model reports the same adverse effect of RI on reading scores for all students. However, the *race-modified* model (Figure 1b) provides the essential context: the RI effect is significantly negative for NH Black students, but not for other race groups. Thus, a race-modified model is necessary to uncover and quantify these racial discrepancies in the effects of structural racism on educational outcomes.

**Fig. 1.**
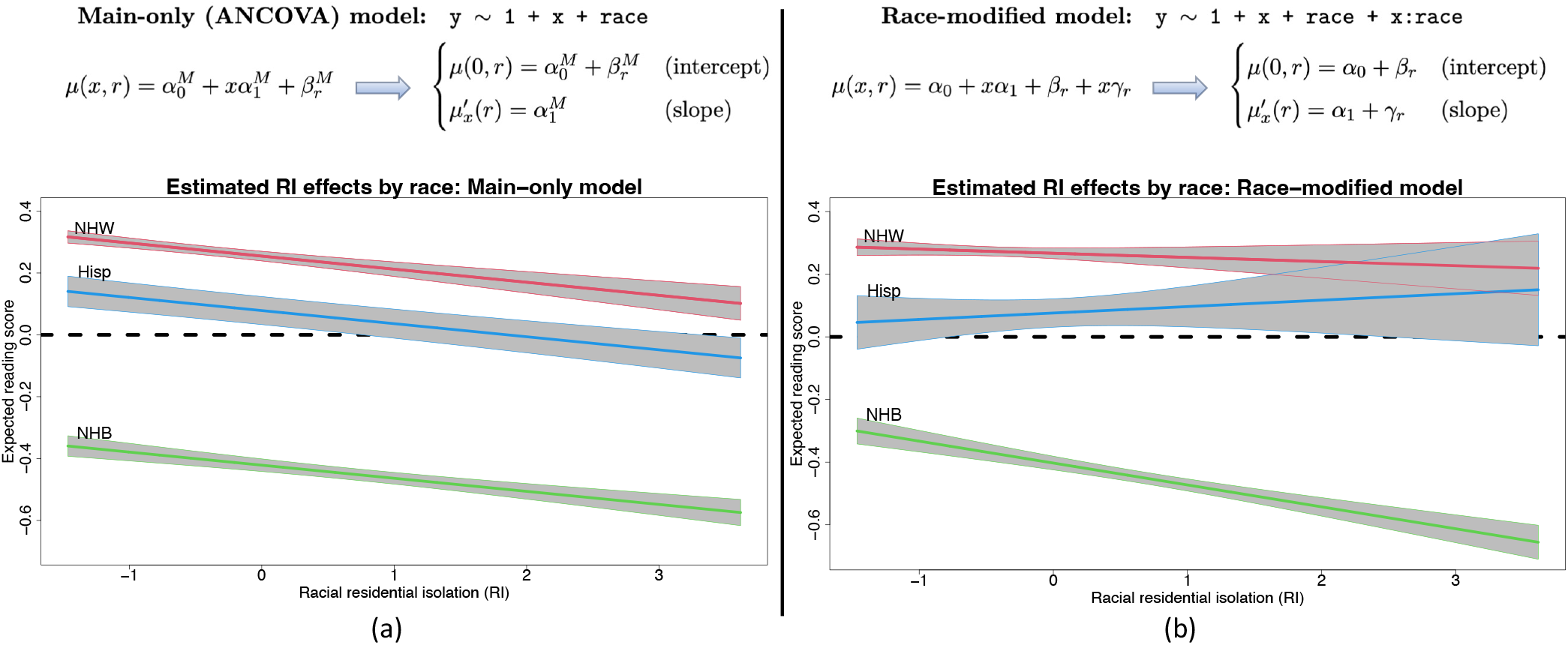
Linear regression models for an outcome variable *Y* with a continuous covariate *X* and a categorical (or nominal) covariate `race`. The models parameterize the expected outcome, 𝔼 (*Y* | *X* = *x*, `race` = *r*) = *µ*(*x, r*), with corresponding *x*-effect (or slope) 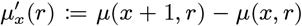. (a) The *main-only* model assumes a global (race-invariant) *x*-effect. (b) The *race-modified* model allows for race-specific *x*-effects. Fit to 4th end-of-grade reading scores in North Carolina, the main-only model obscures important race-specific differences in the effects of racial isolation (RI) that are uncovered by the race-modified model. The negative RI effect observed globally in (a) is driven by the negative RI effect *for non-Hispanic Black (NHB) students*, which does not persist for Hispanic (Hisp) or non-Hispanic White (NHW) students in (b).

Despite these benefits, there are significant racial biases that occur in commonplace estimation, inference, and presentation of results for regression analysis with race as a covariate. It is well-known that both the main-only and race-modified models (Figure 1) are overparametrized: neither {*α*_0_, *β*_*r*_}_*r*_ nor {*α*_1_, *γ*_*r*_}_*r*_ are identifiable without further constraints. Any constant could be added to *α*_0_ and subtracted from each {*β*_*r*_}, and similarly for *α*_1_ and {*γ*_*r*_}, which alters each parameter but leaves the model unchanged. This nonidentifiability is often called the “dummy variable trap” in reference to the use of “dummy variables” to encode categorical variables (Figure 1). Critically, neither the main nor the race-specific parameters can be estimated or interpreted without additional constraints.

Undoubtedly, the most common approach is *reference group encoding* (RGE): a reference group is selected, typically NH White, and removed (*β*_NHW_ = *γ*_NHW_ = 0). This is the default for all major statistical software implementations of (generalized) linear regression, including R, SAS, Python, MATLAB, and Stata, as well as textbook treatments of linear regression (31–33). However, RGE output is racially biased (34), difficult to interpret, and obscures important main and race-modified effects. We categorize these significant limitations into *presentation bias* and *statistical bias*.

## Presentation bias

Table 1 (left) displays standard output for a race-modified model. Under RGE, the RI effect (red) actually refers to the RI effect *only for NH White individuals*, 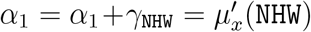. Similarly, Intercept refers to the NH White intercept, *α*_0_ = *α*_0_ +*β*_NHW_ = *µ*(0, `NHW`). We emphasize that the presentation format in Table 1 (left) is predominant in scientific journals. Among recent publications in social science journals, it was found that 92% of such tables used NH White as the reference group, while less than half explicitly stated the reference group (34).

**Table 1.** Linear regression output under default reference group encoding (RGE; left) and *abundance-based constraints* (ABCs; right): race-modified effects of racial isolation (RI) on 4th end-of-grade reading scores for students in North Carolina (y ∼ 1 + RI + race + RI:race).

| Default: reference group encoding (RGE) |  |  | Proposed: abundance-based constraints (ABCs) |  |  |
| --- | --- | --- | --- | --- | --- |
| Variable | Estimate (SE) | p-value | Variable | Estimate (SE) | p-value |
| Intercept | 0.267 (0.009) | <0.001 | Intercept | 0.014 (0.007) | 0.038 |
| Racial isolation (RI) | -0.013 (0.011) | 0.220 | Racial isolation (RI) | -0.032 (0.007) | <0.001 |
| Mother's race |  |  | Mother's race |  |  |
| NH Black | -0.670 (0.014) | <0.001 | NH White | 0.253 (0.006) | <0.001 |
| Hispanic | -0.191 (0.025) | <0.001 | NH Black | -0.417 (0.009) | <0.001 |
| RI × Mother's race | | | Hispanic | 0.062 (0.023) | 0.006 |
| RI:NH Black | -0.057 (0.014) | <0.001 | RI × Mother's race | | |
| RI:Hispanic | 0.034 (0.027) | 0.210 | RI:NH White | 0.018 (0.006) | 0.001 |
|  |  |  | RI:NH Black | -0.038 (0.009) | <0.001 |
|  |  |  | RI:Hispanic | 0.052 (0.024) | 0.031 |

The default regression output (RGE, left) induces *presentation bias*: the estimated Intercept and RI effect (red) refer to the NH White group. This is inequitable, unclear, and misleading: it obfuscates the highly significant and detrimental effects of RI on reading scores (for the main-only model, $\hat{\alpha}_1^M = -0.042$ , $\text{SE}(\hat{\alpha}_1^M) = 0.007$ , $p < 0.001$ ). The regression output under ABCs (right) eliminates presentation bias, confirms the estimated RI effect and standard errors from the main-only model (blue), and clearly highlights and quantifies the critical result that the adverse RI effect is more than doubled for NH Black students

First, this output is *inequitable*: it elevates a single race group above others. Further, all other race-specific effects are presented relative to the NH White group. For instance, `RI:NH Black` refers to the difference between the RI effects for NH Black students and NH White students: 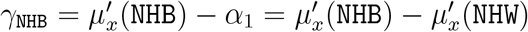. This framing presents NH White as “normal” and other race groups as “deviations from normal”, which is known to bias interpretations of results (35). Second, this output is *unclear* : it is nowhere indicated that the intercept and RI effects are specific to NH White students. A cursory inspection of this output might result in a mistaken interpretation of the RI effect as a global effect, rather than a NH White effect. Finally, this output is *misleading*: the RI effect is reported to be small and insignificant, despite clear evidence to the contrary (Figure 1). Under RGE, the addition of the race-modifier substantially alters the estimates and reduces the statistical power for the RI main effect (*α*_1_).

## Statistical bias

The racial inequity in RGE also permeates statistical estimation and inference. Modern statistical learning commonly features penalized regression, variable selection, and Bayesian inference (36). Broadly, these *regularization* strategies seek to stabilize (i.e., reduce the variance of) estimators, typically by “shrinking” coefficients toward zero. This approach is particularly useful in the presence of a moderate to large number of covariates that may be correlated. However, under RGE, shrinking or setting coefficients to zero introduces racial bias to the estimation. Critically, shrinkage or selection of the race-specific terms, *γ*_*r*_ → 0, does not innocuously shrink toward a global slope; rather, it implies that the coefficient on *x* for race *r* is pulled toward that of the NH White group, 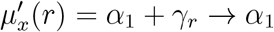, and 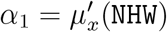 (NHW). Not only is this estimator racially biased, but also it attenuates the estimated differences between the *x*-effects for each race and NH White individuals. Identification and quantification of such race-modified effects are precisely the goals of race-modified models. Furthermore, RGE cannot distinguish between shrinkage toward a global, race-invariant *x*-effect and shrinkage toward the NH White *x*-effect: both require *γ*_*r*_ → 0 for all *r*. A fundamental goal of penalized estimation and selection in this context is to remove unnecessary race-modifiers. However, with RGE, the cost is racial bias in the shrinkage and selection. Thus, default RGE cannot fully and equitably leverage the state-of-the-art in statistical learning.

Although RGE is used in the overwhelming majority of regression analyses, there are several alternatives. *Subgroup analysis* subsets the data by (race) groups and fits separate regression models (12, 25, 30, 37). This approach produces race-specific i ntercepts a nd s lopes, a nd thus implicitly acknowledges the importance of race-modifiers. However, subgroup analysis does not estimate global (race-invariant) *x*-effect estimates or inference, cannot incorporate information-sharing or regularization across race groups (often leading to variance inflation and reduced power), and cannot test for race-modifier effects. *Sum-to-zero* (S TZ) constraints ad dress the in equities in RGE, but the resulting model parameters are difficult to interpret and the estimators do not offer any of the appealing statistical properties provided by our preferred approach. *Overparametrized estimation* omits any identifying constraints and relies on regularized regression to produce unique estimators. But the model parameters remain nonidentified, so the estimates remain extremely difficult to interpret. These estimates also fail to offer the useful statistical properties discussed subsequently. Finally, marginal means (38) use post-processing to provide useful model summaries, and will be identical for RGE, STZ, and ABCs under ordinary least squares estimation. Nonetheless, it remains imperative to choose an identifiable model parametrization that delivers interpretable and equitable model parameters, appealing properties for estimation and inference, and suitable behavior for regularized regression and variable selection. Notably, marginal means that are estimated by regularized regression *will* depend on the model parametrization (RGE, STZ, ABCs, etc.).

The primary goal of this paper is to describe and validate alternative statistical methods that eliminate these racial biases. Our preferred approach ensures equitable and interpretable parameters with accompanying estimators that offer unique and appealing statistical properties. We apply these tools to identify and quantify the race-modified effects of multiple environmental, social, and other factors on 4th end-of-grade readings scores for students in North Carolina. Although we focus on race, the proposed methods remain applicable for other categorical covariates including sex, national origin, religion, and other protected groups. This work is accompanied by a companion paper on statistical theory (39), an `R` package `lmabc` that implements our estimation and inference methods, and an online vignette that provides accessible examples and documentation: https://drkowal.github.io/lmabc/.

## Results

### Abundance-Based Constraints (ABCs) for Linear Regression

We update the race-modified model (Figure 1b) for multivariable regression with *p* covariates ***X*** = (*X*_1_, …, *X*_*p*_)^⊤^, where the effect of each variable may be modified by race:

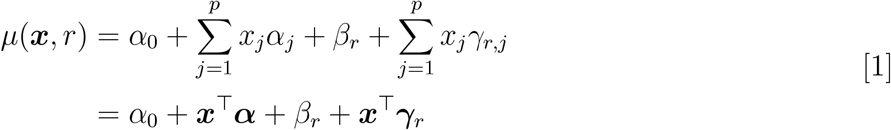

where ***α*** = (*α*_1_, …, *α*_*p*_)^⊤^ are the main *x*-effects a nd ***γ*** _*r*_ = (*γ*_*r*,1_, …, *γ* _*r,p*_)^⊤^ a re t he race-modifier effects. The m ain-only version o mits a ll i nteractions (*γ*_*r,j*_ = 0). The i ntercepts a re race-specific, *µ*(0, *r*) = *α*_0_ +*β*_*r*_, while the race-modified m odel y ields r ace-specific sl opes fo r ea ch variable *j* = 1, …, *p*:

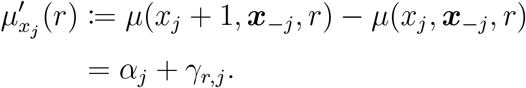

The parameters {*α*_0_, *β*_*r*_}_*r*_ and {*α*_*j*_, *γ*_*r,j*_ }_*r,j*_ must be further constrained to enable unique estimation and meaningful inference. Linear constraints of the form ∑_*r*_ *c*_*r*_*β*_*r*_ = 0 and ∑_*r*_ *c*_*r*_*γ*_*r,j*_ = 0 are most common: RGE sets *c*_1_ = 1 and *c*_*r*_ = 0 for *r >* 1, while STZ uses *c*_*r*_ = 1 for all *r*. However, the equitability, interpretability, and statistical properties of the parameters and estimators depend critically on the choice of {*c*_*r*_}.

We advocate *abundance-based constraints (ABCs)* that use the race group abundances:

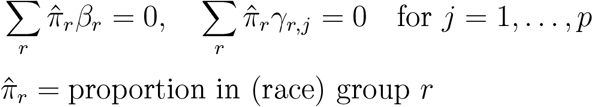

or equivalently, 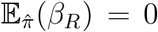 and 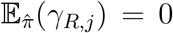 for all *j*, where the expectation is taken `over` a categorical random variable *R* with 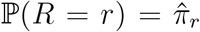. If known, the population proportions may be used for 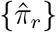; otherwise, we use the sample proportions. ABCs, under various names, have appeared previously, but only for main-only models (40–42). Critically, motivation for this approach is severely lacking; even among previous work that mentions ABCs, they are routinely dismissed in favor of RGE or STZ. Here, we promote ABCs for *race-modified* models based on new arguments for equitability, interpretability, and special statistical properties.

To evaluate equitability and interpretability, we consider the meaning of each parameter in the race-modified model. Under ABCs, the race-modified model satisfies 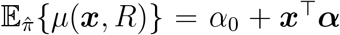, which produces a global (race-invariant) linear regression. As a consequence, each main *x*-effect may be expressed as the *race-averaged slope* for the *j*th variable:

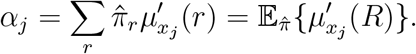

Unlike with RGE, where 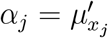 (`NHW`), ABCs do *not* anchor each main *x*-effect to the NH White group and instead provide a global interpretation for these key parameters. The benefits cascade down to the other parameters:

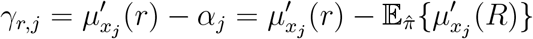

which is the difference between the *race-specific slope* and the *race-averaged slope* for variable *j*. The intercept also retains a convenient, more equitable interpretation. Suppose that each continuous covariate is centered, 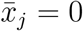. Then the intercept parameter is a marginal expectation:

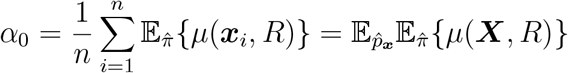

where the expectation is taken (separately) `over` 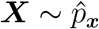 for 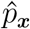 the empirical distribution of 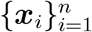 and 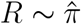. The race-specific intercept coefficients proceed similarly:

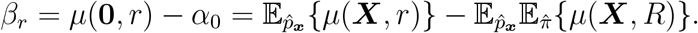

Again, unlike for RGE, the parameters *α*_0_ and *β*_*r*_ no longer elevate the NH White group. Instead, ABCs define all parameters as 1) global, r ace-averaged main effects or 2) race-specific deviations.

### Estimation and Inference

Given data 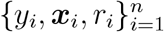, the race-modified m odel w ith A BCs is estimated by applying linearly-constrained ordinary least squares (OLS) estimation. Standard errors, confidence intervals, and hypothesis testing are available as in traditional OLS estimation. Options for regularized (ridge, lasso, etc.) regression are provided (see **Methods**). Because the estimators satisfy ABCs, they retain the same properties and interpretations as the parameters above.

### Statistical Properties

A central obstacle with race-modified models is that, for default approaches (RGE, STZ, etc.), the inclusion of these interaction terms fundamentally alters the interpretations, estimates, and standard errors for the main *x*-effects. We observe this empirically (Table 1, left): compared to the main-only model, the race-modified model *under RGE* attenuates the RI main effect 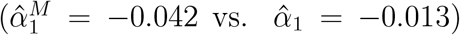 and inflates the standard error 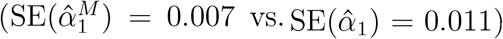. These results are not contradictory: the RI effect is weaker for the NH White group (Figure 1b) than for the aggregate (Figure 1a), while NH White students represent a subset (58%) of the full sample. The broader implication is that analysts may be reluctant to include race-modifiers. However, omitting race-modifiers can produce misleading results (Figure 1).

ABCs resolve these problems. The first key property of ABCs is *estimation invariance*: the OLS estimates of the main *x*-effects are nearly identical between the main-only model and the race-modified model, under appropriate conditions. For *p* = 1 (Figure 1), ABCs uniquely yield the remarkable result

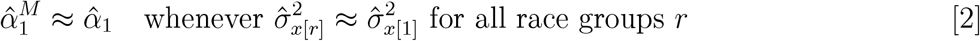

where 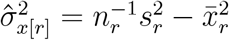 is the (scaled) sample variance of 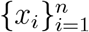 within each race group *r*, with 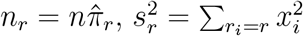 and 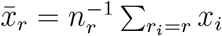. Similar results are available for general *p >* 1 under suitable modifications of the equal-variance condition (39).

The equal-variance condition in Eq. (2) requires that the scale of *x* is approximately the same for each race group. Otherwise, a one-unit change in *x* is not comparable across race groups. In that case, race-specific slopes are necessary, and the global slope from the main-only model 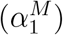 is not a suitable summary. However, the estimation invariance of ABCs is empirically robust to violations of the equal-variance condition. This condition is strongly violated for RI in Table 1 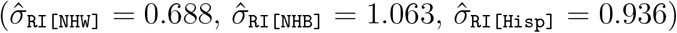, yet the main effect estimates remain similar 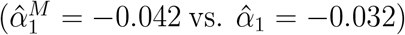 and the standard errors (and *p*-values) are identical 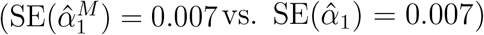. Similar results are observed for simulated data (Figure A.1), which further show that estimation invariance does *not* hold for RGE or STZ.

The second key property of ABCs is related to inference: the main *x*-effect standard errors are *equal or smaller* under the race-modified model, 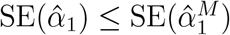, whenever 1) the equal-variance condition in Eq. (2) holds and 2) the estimated residual variance is equal or smaller under the race-modified model than the main-only model (39). When the race-modifier term (x:race) has small or moderate impact, then the standard errors for the main *x*-effect are approximately the same between the main-only and race-modified models (see Table 1 and Figure 2). However, if the race-modifier term explains substantial variability in *Y*, then the race-modified model can actually *increase statistical power* for the main *x*-effect compared to the main-only model. Thus, contrary to intuition, the race-modified model—with its greater complexity and additional parameters to estimate—provides superior inference for the main *x*-effect than in the simpler, main-only model.

**Fig. 2.**
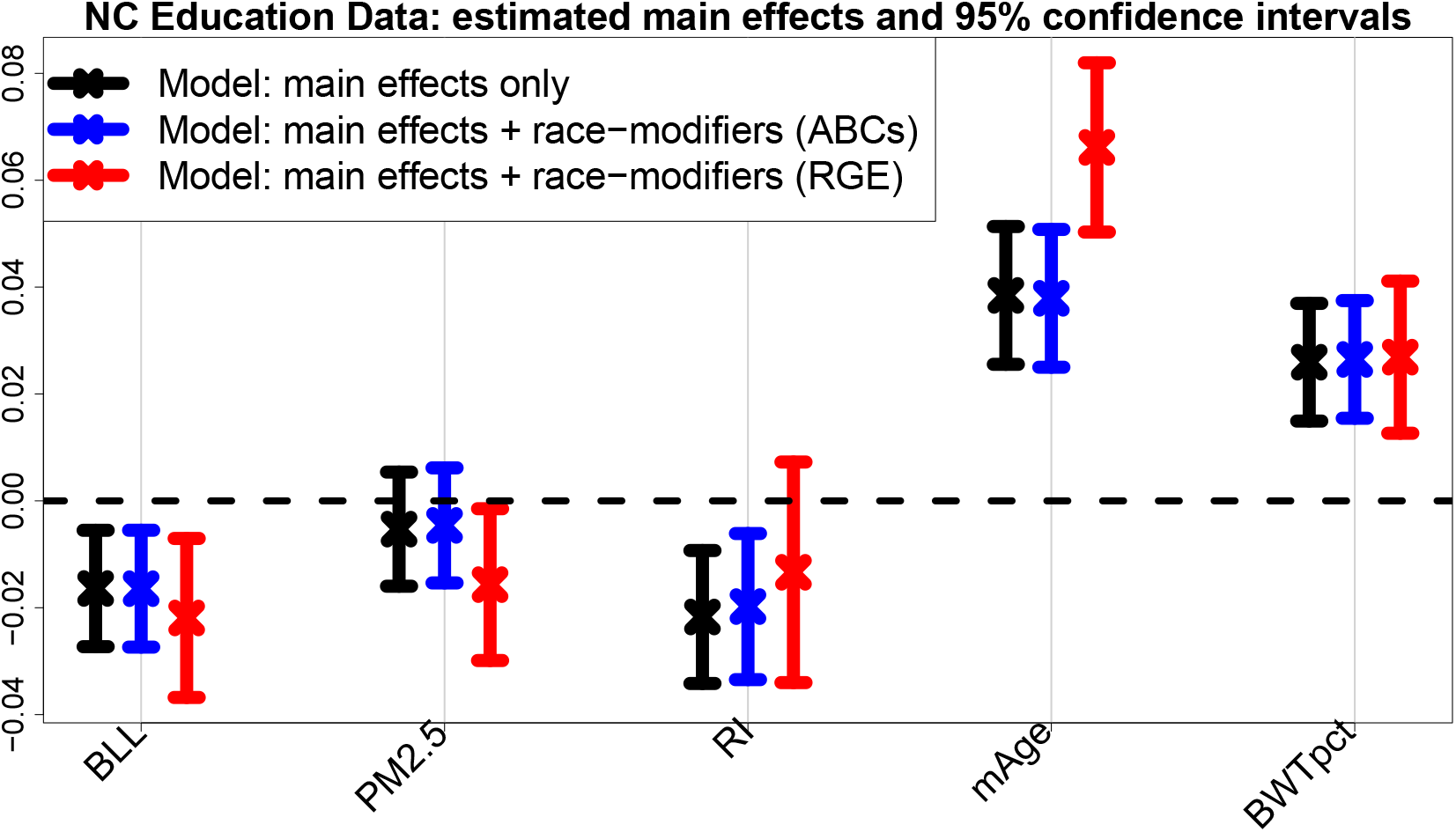
Estimates and 95% confidence intervals for the main effects in the multivariable regression *without* race-modifiers (black) and the multivariable regression *with* race-modifiers under ABCs (blue) and RGE (red). Results are presented for blood lead level (BLL), PM_2.5_ exposure (PM_2.5_), racial isolation (RI), mother’s age (mAge), and birthweight percentile for gestational age (BWTpct), each of which is interacted with race in the expanded model (blue, red); additional covariates include sex, mother’s education level, mother’s marital status, mother’s smoking status, and economically disadvantaged (Table 2). ABCs exhibit invariance: despite the additional race-modifier parameters, the point and interval estimates for the main effects (blue) are nearly indistinguishable from those in the main effects-only model (black), thus effectively allowing the inclusion of race-modifiers “for free”. In contrast, the RGE terms (red) correspond to the *x*-effects for the NH White group and deviate substantially for PM_2.5_, RI, and mAge, including shifts in location and much wider intervals.

With ABCs, the analyst may include race-modifiers “for free”: the estimates and inference for the main *x*-effects are nearly unchanged by the addition of race-modifiers (x:race). This result is unique to ABCs and makes no assumptions about the true relationship between *Y, X*, and race. Notably, arbitrary dependencies are permitted between *X* and race—including varying means and distributions of *X* by race group—as long as the equal-variance condition holds. Thus, this result is distinct from classical estimation invariance results with OLS that require uncorrelatedness (43).

**Table 2.** Linear regression output (under ABCs) for the race-modified effects of environmental, social, and other factors on 4th end-of-grade reading scores for students in North Carolina.

| Variable | Estimate (SE) | p-value | Variable (continued) | Estimate (SE) | p-value |
| --- | --- | --- | --- | --- | --- |
| Intercept | 0.000 (0.007) | 0.976 | BLL × Mother's race | | |
| Blood lead level (BLL) | -0.016 (0.006) | 0.003 | BLL:NH White | -0.005 (0.005) | 0.240 |
| PM<sub>2.5</sub> exposure (PM<sub>2.5</sub>) | -0.005 (0.005) | 0.403 | BLL:NH Black | 0.002 (0.007) | 0.792 |
| Racial isolation (RI) | -0.020 (0.007) | 0.005 | BLL:Hispanic | 0.042 (0.022) | 0.060 |
| Mother's age (mAge) | 0.038 (0.007) | <0.001 | PM<sub>2.5</sub> × Mother's race | | |
| Birthweight percentile for gestational age (BWTpct) | 0.026 (0.006) | <0.001 | PM<sub>2.5</sub>:NH White | -0.011 (0.005) | 0.018 |
| Mother's race |  |  | PM<sub>2.5</sub>:NH Black | 0.021 (0.007) | 0.005 |
| NH White | 0.173 (0.006) | <0.001 | PM<sub>2.5</sub>:Hispanic | -0.017 (0.023) | 0.472 |
| NH Black | -0.320 (0.010) | <0.001 | RI × Mother's race | | |
| Hispanic | 0.258 (0.024) | <0.001 | RI:NH White | 0.006 (0.006) | 0.250 |
| Sex |  |  | RI:NH Black | -0.020 (0.008) | 0.016 |
| Male | -0.064 (0.005) | <0.001 | RI:Hispanic | 0.058 (0.023) | 0.013 |
| Female | 0.064 (0.005) | <0.001 | mAge × Mother's race | | |
| Mother's education level |  |  | mAge:NH White | 0.028 (0.005) | <0.001 |
| Did not complete high school | -0.179 (0.011) | <0.001 | mAge:NH Black | -0.040 (0.007) | <0.001 |
| Completed high school | -0.074 (0.007) | <0.001 | mAge:Hispanic | -0.032 (0.024) | 0.193 |
| At least some postsecondary | 0.181 (0.008) | <0.001 | BWTpct × Mother's race | | |
| Mother's marital status |  |  | BWTpct:NH White | 0.000 (0.005) | 0.929 |
| Married at time of birth | 0.018 (0.006) | 0.003 | BWTpct:NH Black | 0.002 (0.007) | 0.766 |
| Not married at time of birth | -0.024 (0.008) | 0.003 | BWTpct:Hispanic | -0.018 (0.022) | 0.427 |
| Mother's smoking status |  |  |  |  |  |
| Smoker | -0.039 (0.013) | 0.003 |  |  |  |
| Non-smoker | 0.008 (0.003) | 0.003 |  |  |  |
| Economically disadvantaged |  |  |  |  |  |
| Yes | -0.109 (0.005) | <0.001 |  |  |  |
| No | 0.171 (0.009) | <0.001 |  |  |  |

Data restricted to individuals with 37-42 weeks gestation, mother's age 15-44 years old at birth, BLL $\leq 80\mu\text{g}/\text{dL}$ (and capped at $10\mu\text{g}/\text{dL}$ ), birth order $\leq 4$ , no current English language learners, and residence in NC at the time of birth and time of 4th end-of-grade test. "Economically disadvantaged" is determined by participation in the National Lunch Program.

### Sparsity

Sparsity is often prioritized to remove extraneous parameters, reduce estimation variability, and simplify interpretations. Regularized regression can produce sparse estimates, but depends critically on the parametrization. Importantly, sparsity of the race-modifiers, *γ*_*r,j*_ = 0, is meaningful under ABCs: it implies that the race-specific slope equals the race-averaged slope, 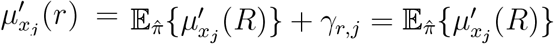. This eliminates the racial bias and inequity under RGE, where the same sparsity instead implies that the race-specific slope equals the NH White slope, 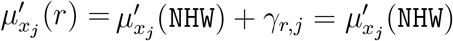.

An especially concerning case arises when the race-modifier is nonzero (*γ*_*r,j*_ ≠ 0), but the main *x*-effect is zero (*α*_*j*_ = 0). Statistical approaches often eschew this scenario, and instead require that interactions are nonzero *only if* a main effect is nonzero (44, 45). Such restrictions are not necessary for ABCs: it is plausible that some race-specific *x*-effects are nonzero, 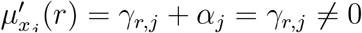, while the race-averaged *x*-effect is zero, 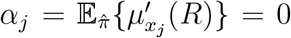. Alarmingly, fitting a main-only model would produce misleading results. Applying Eq. (2), the estimated *x*-effect would be near zero, 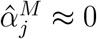, when in fact the *x*-effect is both *significant* and *race-specific*. Thus, it is possible that existing quantitative analyses based on main-only models (i.e., without race-modifiers) obscure both important and race-specific effects of certain variables (Figures 1 and 2).

## NC Education Data Analysis

We apply the proposed methods to study the effects of multiple environmental, social, and other factors on educational outcomes—and assess whether, and how, these effects vary by race. Using ABCs, we fit equitable and interpretable race-modified models, empirically evaluate estimation and inference invariance properties, and study regularized (lasso) regression solution paths under competing parametrizations.

### Data overview

We construct a cohort of *n* = 27, 638 students in North Carolina (NC) by linking three administrative datasets:

*NC Detailed Birth Records* include maternal and infant characteristics for all documented live births in NC. We compute maternal covariates—mother’s race, age (mAge), education level, marital status, and smoking status—and child covariates, sex and birthweight percentile for gestational age (BWTpct). RI is computed using residential addresses at birth.

*NC Blood Lead Surveillance* includes blood lead level (BLL) measurements for each child. Lead is an adverse environmental exposure with well-known effects on cognitive development and educational outcomes (46, 47).

*NC Standardized Testing Data* contains 4th end-of-grade standardized reading scores, economic disadvantage status (determined by participation in the National Lunch Program), and residential address at time-of-test. The reading scores, standardized by the year of test (2010, 2011, or 2012), serve as the outcome variable *Y*. The residential information is used to estimate the average PM_2.5_ exposure (PM_2.5_) `over` the year prior to the test, which is an adverse environmental exposure linked to educational outcomes (48).

Data characteristics are in Table A.1; additional details are provided elsewhere (30, 49, 50). Data management, access, and analysis are governed by data use agreements and an Institutional Review Board–approved research protocol at the University of Illinois Chicago.

### Race-modified regression with ABCs

We estimate a multivariable linear regression for 4th end-of-grade reading scores that includes these environmental, social, and other factors, as well as race-modifiers (Table 2). Each continuous covariate (BLL, PM_2.5_, RI, mAge, and BWTpct) is centered and scaled and each categorical variable (mother’s race, child’s sex, mother’s education level, mother’s marital status, mother’s smoking status, and economically disadvantaged) is identified using ABCs (see **Methods**). Race-modifiers are included for BLL, PM_2.5_, RI, mAge, and BWTpct. Standard model diagnostics confirm linearity, homoskedasticity, and Gaussian error assumptions.

ABCs generate output for all main effects, all race-modifier effects, and each group in every categorical variable, which eliminates the presentation bias that would otherwise accompany *each* categorical variable under RGE. There are highly significant (*p <* 0.01) negative effects for BLL and RI, where the adverse RI effect *doubles* for NH Black students 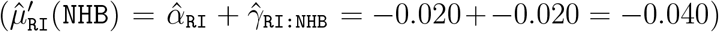. This critical result for RI expands upon the previous model fit (Table 1): here, the model adjusts for many additional factors, yet the effect persists. Significantly lower test scores also occur for students who are NH Black, Male, or economically disadvantaged, and whose mothers who are less educated, unmarried, or smokers at time of birth. Significant positive effects are observed for the opposite categories—which is a byproduct of ABCs (e.g., the Male and Female proportions are identical, so the estimated effects must be equal and opposite)—as well as mAge and BWTpct. Finally, PM_2.5_ is not identified as a significant main effect (*p* = 0.403), yet the race-specific effects *are* significant. Alarmingly, a fitted main-only model (*without* race-modifiers) conveys an insignificant PM_2.5_ effect (Figure 2), which oversimplifies and misleads.

### Estimation invariance with ABCs

Figure 2 presents the estimates and 95% confidence intervals for the main effects that are modified by race. We compare the main-only model (variables only in the left column of Table 2) to the race-modified model (all terms in Table 2), including both ABCs and RGE output for the race-modified models. Remarkably, under ABCs, the estimates and uncertainty quantification for the simpler, main-only model are nearly indistinguishable from those for the expanded, race-modified model. Evidently, ABCs allow estimation and inference for numerous race-specific effects (Table 2, right column) “for free”: the inferential summaries for the main effects are unchanged by the expansion of the model to include race-modifiers. This result empirically confirms the multivariable extension of Eq. (2), despite moderate violations of the equal-variance condition (Table A.2). Unsurprisingly, no such invariance holds for RGE (red): the point and interval estimates are substantially different, with uniformly wider intervals and conflicting conclusions about nonzero coefficients (PM_2.5_, RI). These concerning discrepancies occur because the RGE “main effects” are exclusively for NH White students.

### Regularized regression with ABCs

We assess regularized regression and variable selection with ABCs using lasso regression, including all variables from Table 2. We report estimates across tuning parameter values *λ* for the model coefficients 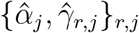 and the race-specific slopes 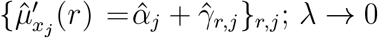 yields OLS estimates, while *λ* → ∞ yields sparse estimates. Since the penalized estimates depend critically on the parameterization, we compare ABCs and RGE. The estimated *λ*-paths for RI are in Figure 3; results for the remaining race-modified effects (BLL, PM_2.5_, mAge, and BWTpct) are in Figures A.2–A.5. RGE fixes 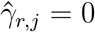 for all *λ*, which results in 1) racially-biased shrinkage of the race-specific effects toward the NH White-specific effect and 2) attenuation of the RI effect 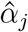 (Figure 3, top right). ABCs resolve these issues. First, the model parameters are separately and equitably pulled toward zero (Figure 3, top left). Second, the RI effect 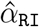 is *not* attenuated, and preserves its magnitude until log *λ* ≈ 5 (Figure 3, top left). Finally, the race-specific RI effects merge at a *global*, and negative, RI effect estimate, and this variable is selected by the one-standard-error rule (36) for choosing *λ* (Figure 3, bottom left).

**Fig. 3.**
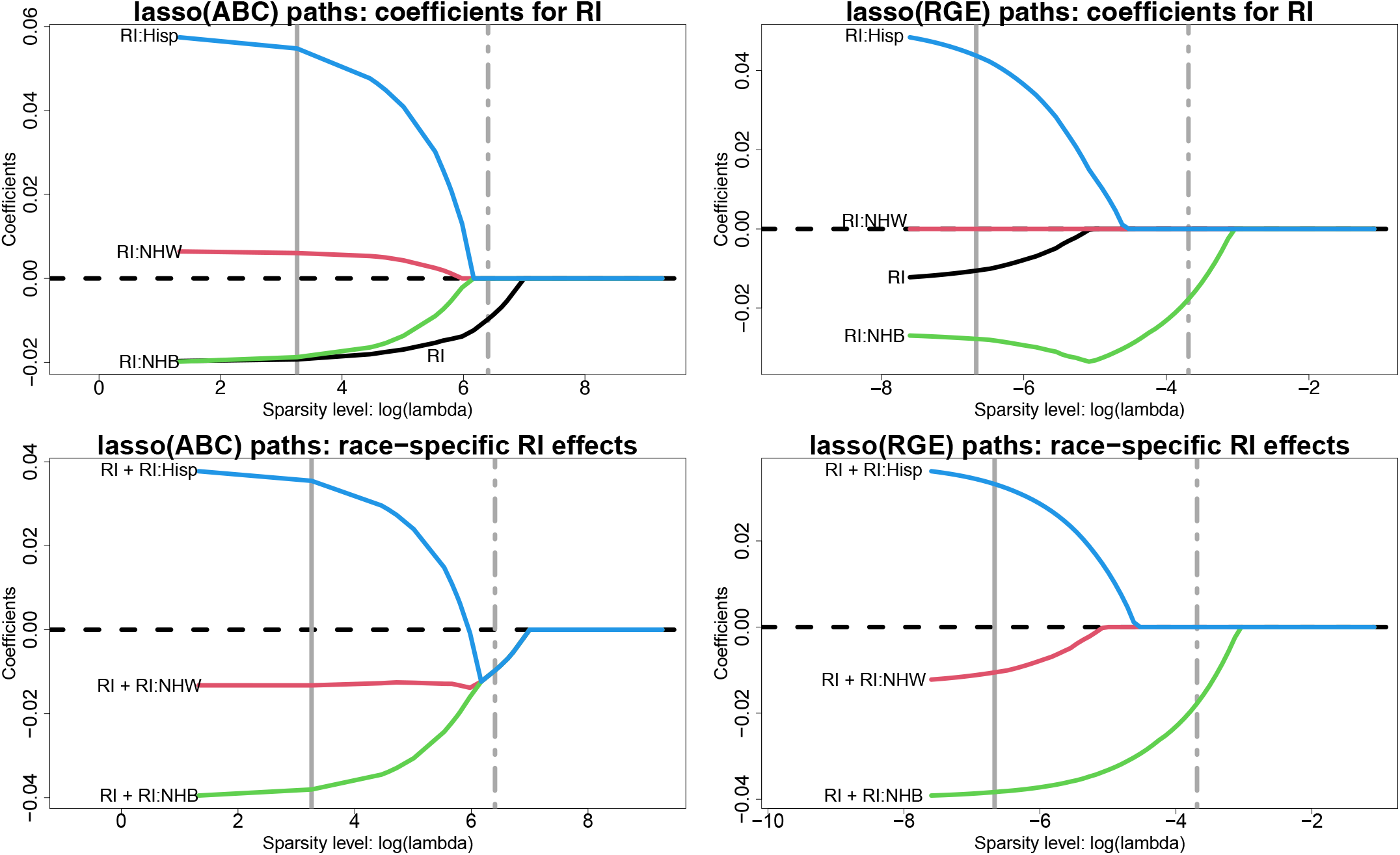
Estimated lasso paths for RI across varying sparsity levels (log *λ*) for the model coefficients 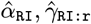 (top) and the race-specific slopes 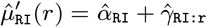 (bottom) under ABCs (left) or RGE (right); vertical lines identify *λ* for the minimum cross-validated error (solid) and one-standard-error rule (dot-dashed). The outcome is 4th end-of-grade reading score and the covariates include all variables in Table 2. Small *λ* approximately corresponds to OLS, while increasing *λ* yields sparsity. Under RGE, the estimates are pulled toward the reference (NH White) estimate—inducing *statistical bias* by race—and the RI effect is attenuated. By comparison, ABCs offer more equitable shrinkage toward a global RI effect, which is nonzero and detrimental for 4th end-of-grade reading scores.

These themes persist for the remaining race-modified effects (Figures A.2–A.5). We supplement the ABC and RGE lasso paths by including the lasso paths for *overparametrized estimation* (Over), which does not include any identifiability constraints. The parameters cannot be estimated uniquely by OLS, but can be estimated by lasso regression with *λ >* 0. In most cases, `Over` sets one of the coefficients {*α*_*j*_, *γ*_*r,j*_}_*r*_ to zero immediately (small *λ*) for each variable *j*. This effect reproduces RGE and thus `Over` inherits the same racial biases in estimation and selection. When this implicit selection sets 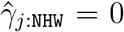, then the `Over` paths resemble those for RGE (RI, not shown; BWTpct, Figure A.5); when the selection corresponds to the smallest 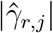 among race groups *r* from ABCs, then the `Over` and ABC paths are similar (BLL, Figure A.2; BWTpct, Figure A.5). However, when this selection sets the main effect to zero, 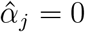 (PM_2.5_, Figure A.3) or overshrinks multiple coefficients toward zero (mAge, Figure A.4), then the `Over` paths differ substantially from both the RGE and ABC paths and demonstrate erratic behavior (Figure A.4).

## Estimation and predictive accuracy for simulated data

We evaluate estimation and prediction for ABCs, RGE, and `Over` across several estimation methods: OLS, ridge, and lasso regression (Figure 4). Data are simulated from a Gaussian main-only model with *p* = 10 covariates and a categorical variable with four levels. For fair comparisons, the data-generating process satisfies both RGE and ABCs. To mimic the challenges of real data analysis, the fitted models are misspecified as Eq. (1), and thus contain extraneous race-modifiers. Root mean squared errors are computed for the regression coefficients {*α*_0_, *α*_*j*_, *β*_*r*_, *γ*_*r,j*_}_*r,j*_, the race-specific slopes {*α*_*j*_ + *γ*_*r,j*_}_*r,j*_, and the model expectations *µ*(***x***, *r*) across 500 simulated datasets. In each case, ABCs are substantially more accurate within each estimation method (OLS, ridge, lasso). The estimation invariance of ABCs offers a plausible explanation: whereas each fitted model includes extraneous variables (the race-modifiers), only ABCs reproduce the main effect estimates from the main-only model, which here is the ground truth. This unique statistical property of ABCs is not only convenient for interpreting race-modified models, but also provides more accurate estimates and predictions under both OLS and regularized regression.

**Fig. 4.**
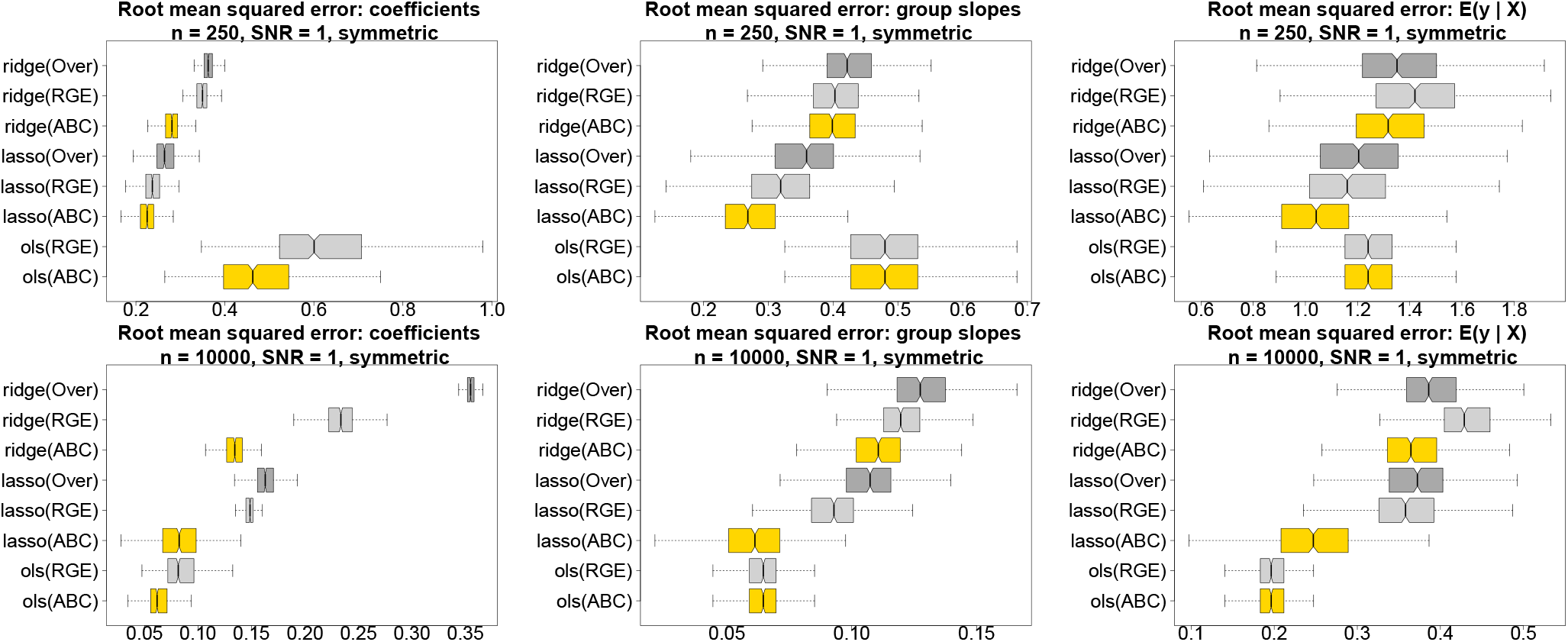
Estimation and prediction accuracy for the regression coefficients (left), the race-specific slopes (center), and the fitted values (right) for *n* = 250 (top) and *n* = 10,000 (bottom) across 500 simulated datasets; nonoverlapping notches indicate significant differences between medians. Data are generated from a Gaussian main-only model with *p* = 10 covariates and a categorical variable with symmetric proportions ***π*** = (0.15, 0.35, 0.15, 0.35)^⊤^; both RGE and ABCs are satisfied in the true data-generating process. All fitted models use the race-modified model Eq. (1) and thus contain extraneous race-modifiers. ABCs (gold) outperform both RGE (light gray) and `Over` (dark gray) within each estimation method (ridge, lasso, OLS). By definition, the OLS race-specific slopes and fitted values are invariant to the constraints (ABCs or RGE), and `Over` cannot be computed for OLS.

## Discussion

The path to more equitable decision-making and policy requires a precise and comprehensive understanding of the links between race and health and life outcomes. Alarmingly, the primary statistical tool for this task—regression analysis with race as a covariate and a modifier—in its current form propagates racial bias in both the *presentation* of results and the *estimation* of model parameters. We advocated an alternative approach, abundance-based constraints (ABCs), with several unique benefits. First, ABCs eliminate these racial biases in both presentation and statistical estimation of linear regression models. Second, ABCs produce more interpretable parameters for race-modified models. Third, estimation with ABCs features an appealing invariance property: the estimated main effects are approximately unchanged by the inclusion of race-modifiers. Thus, analysts can include and estimate race-specific effects “for free”—without sacrificing parameter interpretability, equitability, or statistical efficiency. Finally, ABCs are especially convenient for regularized regression and variable selection, with meaningful and equitable notions of parameter sparsity and efficient computational algorithms.

Using this new approach, we estimated the effects of multiple environmental, social, and other factors on 4th end-of-grade readings scores for a large cohort of students (*n* = 27, 638) in North Carolina. In aggregate, this analysis 1) identified significant race-specific effects for racial (residential) isolation, PM_2.5_ exposure, and mother’s age at birth; 2) showcased the racial biases and potentially misleading results obtained under default approaches; and 3) provided more equitable and interpretable estimates, uncertainty quantification, and selection, both for main effects and race-modified effects. Simulation studies demonstrated substantially more accurate estimates and predictions with OLS, ridge, and lasso regression compared to alternative approaches.

We acknowledge that the interpretation of any “race” effect requires great care (51). Race encompasses a vast array of social and cultural factors and life experiences, with effects that vary across time and geography (27, 52). In some settings, race data are unreliable or partially missing (53, 54). These overarching challenges are not addressed in this paper.

## Methods

### Abundance-Based Constraints (ABCs) with Multiple Categorical Covariates

Regression analysis often features multiple continuous covariates ***X*** = (*X*_1_, …, *X*_*p*_)^⊤^ *and* multiple categorical covariates ***R*** = (*R*_1_, …, *R*_*L*_)^⊤^ such as race, sex, education level, etc.:

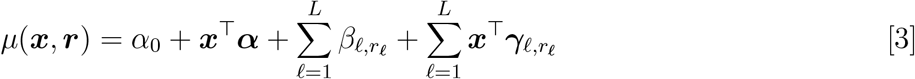

where ***α*** = (*α*_1_, …, *α*_*p*_)^⊤^ and 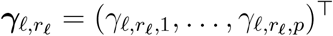 are *p*-dimensional and *r*_𝓁_ denotes the level of the 𝓁th categorical variable, 𝓁 = 1, …, *L*. Eq. (3) includes all continuous-categorical interactions and requires *L*(1 + *p*) constraints for identification; RGE sets *β*_𝓁,1_ = 0 for all 𝓁 and *γ*_𝓁,1,*j*_ = 0 for all 𝓁, *j*.

We extend the definition of ABCs based on the *joint* distribution of the categorical variables ***R***. Specifically, let 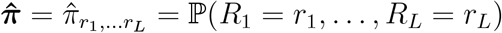. If known, the population proportions may be used for 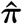; otherwise, we use the sample proportions based on the observed data 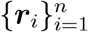, i.e., 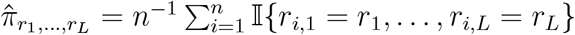. Concisely, the generalized ABCs are

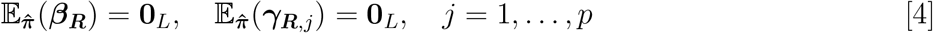

where 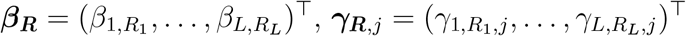, and 0_*L*_ is an *L*-dimensional vector of zeros. Eq. (4) may be equivalently represented via separate marginal expectations for the *L* sets of categorical covariate parameters: for instance, 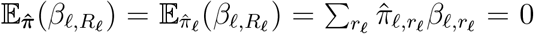 for each categorical covariate 𝓁.

ABCs in Eq. (4) provide interpretable parameter identifications with equitable presentation and estimation. These interpretations are unchanged if some or all interaction terms are omitted from Eq. (3), which may occur if multiple categorical variables (e.g., sex, education level) are included as covariates, but only race is included as a modifier. ABCs imply that 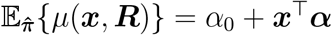, so that averaging the regression Eq. (3) `over` all categorical variables (jointly) yields a multivariate regression with only continuous variables. Individually, each *x*_*j*_-effect satisfies

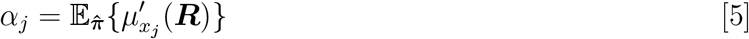

where 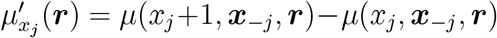 is the slope in the *j*th direction. To further simplify the interpretation, the expectation under 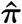 in Eq. (5) need only be taken with respect to the categorical variables that are *interacted* with *x*_*j*_ (e.g., race). By comparison, the RGE parametrization yields 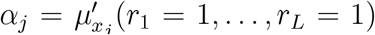, which is the group-specific slope for *x*_*j*_ with each group set to its reference category (e.g., NH White, Male, etc.). Clearly, this representation compounds inequity across each categorical variable and fails to deliver a global interpretation of the *x*_*j*_-effect.

Interpretation of group-specific slopes and the parameters *γ*_𝓁,*r, j*_ proceeds by considering *partial* expectations 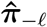, which is analogous to the joint distribution 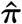 but omits the 𝓁th categorical variable. Here, as with Eq. (5), this expectation need only consider the categorical variables that are interacted with *x*_*j*_; if the 𝓁th categorical variable is the only interaction term, then no expectation is needed at all. Then the *x*_*j*_-effect when the 𝓁th categorical variable has level *r*_𝓁_, averaged `over` the remaining categorical variables, is

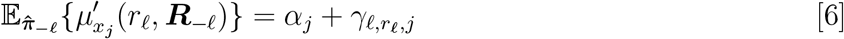

or equivalently, 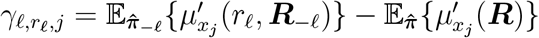. The interpretation is simpler than the notation: Eq. (6) directly extends the usual notion of race-specific slopes to average `over` any other categorical variables that modify *x*_*j*_.

### Estimation

Statistical estimation with ABCs requires solving a linearly-constrained least squares problem given data 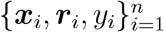. Define ***θ*** to be the model parameters 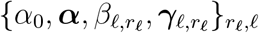 and 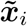 to include the intercept, covariates, race variable indicators (i.e., “dummy variables”), and covariate-race interactions such that Eq. (3) may be written 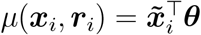. Let ***C*** encode ABCs such that ***Cθ*** = 0 enforces Eq. (4), so ***C*** has *m* = *L*(1 + *p*) rows corresponding to the number of constraints. The OLS estimator under ABCs is

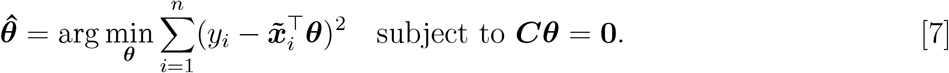

To compute 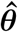 —and subsequently provide inference and penalized estimation—we reparametrize the problem into an unconstrained space with *m* fewer parameters. Let ***C***^⊤^ = ***QR*** be the QR-decomposition of the transposed constraint matrix with columnwise partitioning of the orthogonal matrix ***Q*** = (***Q***_1:*m*_ : ***Q***_−(1:*m*)_) with ***R***^⊤^ = (***R***_1:*m*,1:*m*_ : 0), since ***C***^⊤^ has rank *m*. It is straightforward to verify that ***θ*** = ***Q***_−(1:*m*)_***ζ*** satisfies ***Cθ*** = 0 for *any* ***ζ***. Then, using the adjusted covariate matrix 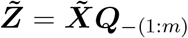 with 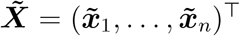, the solution to Eq. (7) is equivalently solved using unconstrained OLS:

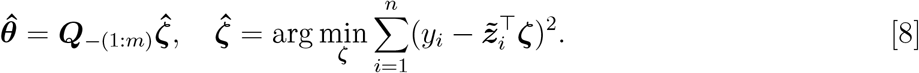

The QR-decomposition has minimal cost due to the efficiency of Householder rotations and the low dimensionality of ***C*** (55).

Although alternative computing strategies are available, the reparametrization in Eq. (8) is especially convenient for generalizations to regularized (lasso, ridge, etc.) estimation. Let 𝒫 (***θ***) denote a complexity penalty on the regression coefficients. The penalized least squares estimator under ABCs is

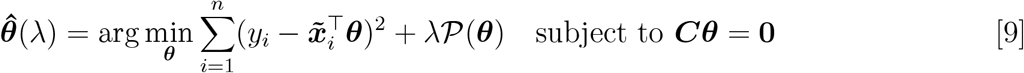

where *λ* ≥ 0 controls the tradeoff between goodness-of-fit and complexity (measured via 𝒫). Following Eq. (8), we instead compute

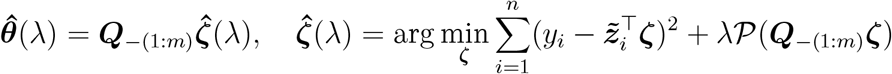

which requires the solution to an *unconstrained* penalized least squares problem.

We focus on complexity penalties of the form

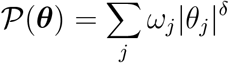

where *ω*_*j*_ *>* 0 are known weights, *δ* = 1 produces sparse coefficients (a daptive la sso re gression) and *δ* = 2 guards against collinearity (adaptive ridge regression). Under ridge regression (*δ* = 2), the solution is

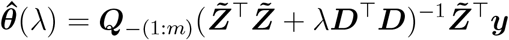

where 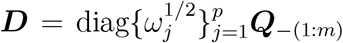. The lasso version (*δ* = 1) can be solved efficiently using the genlasso package in R (56).

For practical use, we set *ω*_*j*_ to be the sample standard deviation of the *j*th column of 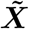 (with *ω*_1_ = 1 for the intercept). This strategy applies a standardized penalty to each covariate, which is especially important for ABCs. In particular, the magnitudes of the race-specific coefficients vary according to the abundance of the group: by construction, low abundances in group *r* will correspond to larger group *r*-specific coefficients. The standardized penalty adjusts for this effect to avoid overpenalization of group-specific coefficients for groups with low abundance.

### Inference

The reparametrization strategy in Eq. (8) allows direct application of classical inference theory to the ABC OLS estimator: 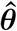 is a known, linear function of the (unconstrained) OLS estimator 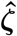. Thus, it is straightforward to derive the (Gaussian) sampling distribution of the ABC OLS estimator, which can be used to compute standard errors, hypothesis tests, and confidence intervals, and to establish unbiasedness and efficiency of the estimator. Under minimal assumptions, the (unconstrained) OLS estimator satisfies 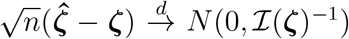 and thus the ABC OLS estimator satisfies

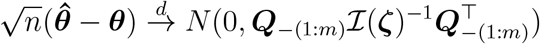

where ℐ is the Fisher information and ***ζ, θ*** are the true parameter values. When the regression model is paired with independent and identically distributed Gaussian errors *ϵ*_*i*_ := *y*_*i*_ − *µ*(***x***_*i*_, ***r***_*i*_) with variance *σ*^2^, the unconstrained OLS estimator satisfies 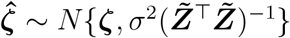 and thus

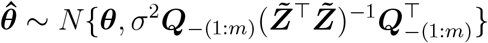

even infinite samples. This sampling distribution for the OLS estimator under ABCs ensures unbiasedness and efficiency, and provides the means to compute standard errors, hypothesis tests, and confidence intervals.

### Simulation design

Each simulated dataset is constructed from a Gaussian multivariable main-only model

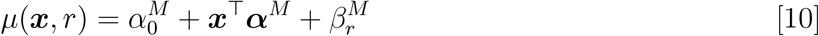

with *p* = 10 continuous covariates and one categorical (race) covariate with four levels. The *p* = 10 continuous covariates include six independent covariates, *X*_*j*_ ∼ *N* (0, 1) for *j* = 1, 2, 3, 6, 7, 8, and four covariates that depend on the categorical variable, [*X*_*j*_ | *R* = *r*] ∼ *N* (*r*, 1), i.e., mean one for group one, mean two for group two, etc., for *j* = 4, 5, 9, 10. The categorical variable is generated based on population proportions ***π***, which we describe below. In addition to the intercept *α*_0_ = 1, the true coefficients for the continuous covariates are *α*_1_ = … = *α*_5_ = 1 (signals) and *α*_6_ = … = *α*_10_ = 0 (noise). Only main effects (i.e., no race-modifiers or interactions) are included in the data-generating process. We vary the sample size, *n* ∈ {250, 10,000}, and use a signal-to-noise ratio of one.

The categorical covariate and accompanying coefficients are constructed carefully to ensure fair comparisons. Different identifications correspond to different parameterizations, which may lead to unfair evaluations. Thus, we use a data-generating model that satisfies both RGE and ABCs. We consider two designs for the categorical proportions: symmetric weights ***π*** = (0.15, 0.35, 0.15, 0.35)^⊤^ (Figure 4) and uniform weights ***π*** = (0.25, 0.25, 0.25, 0.25)^⊤^ (Figure A.6). The true categorical variable coefficients are ***β*** = (0, 1, 0, −1)^⊤^, so RGE is satisfied, *β*_1_ = 0. ABCs are satisfied for the true population proportions, ∑ _*r*_ *π*_*r*_*β*_*r*_ = 0, but we use the sample proportions 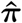 for estimation. The omission of race-modifiers from the data-generating process satisfies both RGE and ABCs. Finally, we require at least *p* + 1 observations for each categorical level, which is necessary for OLS estimation of the interaction effects. We simulate 500 such datasets for each (*n*, ***π***) design.

All competing methods follow Eq. (1), which includes all continuous covariates, race, and all race-modifiers. Thus, all competing models are overparametrized relative to the ground truth, with 55 columns of the unconstrained designed matrix 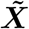 and 44 identifiable model parameters to estimate. The estimation approaches are OLS, ridge regression, and lasso regression. The tuning parameter for ridge and lasso regression is selected using the one-standard-error rule (36). The parametrizations determine the identification constraints on *β*_*r*_ and *γ*_*r,j*_: we consider ABCs, RGE, and Over. `Over` is not identified for OLS, and thus is presented only for ridge and lasso regression.

### Empirical verification of estimation invariance with ABCs

To empirically verify estimation invariance with ABCs, we generate 500 synthetic datasets that mildly violate the equal-variance condition. Iteratively, we sample a categorical variable *R* with groups {`A, B, C, D`} and respective probabilities ***π*** = (0.55, 0.20, 0.10, 0.15)^⊤^ and then sample a continuous variable with the distribution determined by the group:

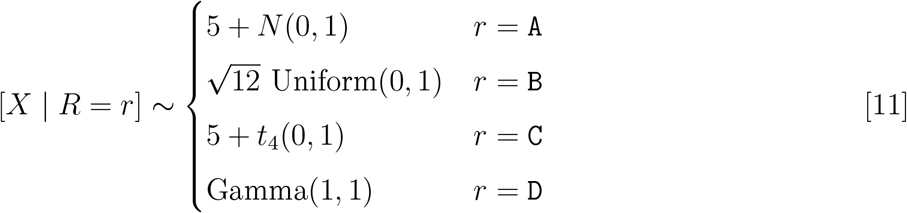

By design, *X* depends on *R* in both mean and distribution. The *R*-specific *population* variances of *X* are each one, but Eq. (2) requires that the *R*-specific *sample* variances are identical, which will not be satisfied for any simulated dataset. Thus, Eq. (11) includes a mild deviation from the equal-variance condition Eq. (2).

The response variable *Y* is simulated with expectation

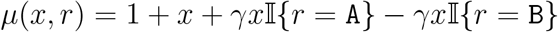

plus *t*_4_(0, 1)-distributed errors, i.e., standard *t*-distributions with 4 degrees of freedom. The coefficient *γ* determines the strength of the race-modifier e ffect: we co nsider *γ* = 0 (no race-modifier effect), *γ* = 0.5 (50% of the main *x*-effect), and *γ* = 1.5 (150% of the main *x*-effect). This data-generating process includes a *R*-modifier (when *γ* ≠ 0) but does not satisfy ABCs or traditional Gaussian error assumptions. Repeating this process 500 times, each simulated dataset contains 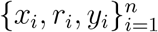. We consider *n* ∈ {100, 500}.

We fit the main-only and race-modified models and record the estimated *x*-effects 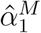 and 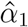, respectively, for each simulated dataset. These estimated coefficients depend on the constraints: we compare ABCs, RGE (with reference group A), and STZ constraints for {*β*_*r*_, *γ*_*r*_} (Figure A.1). Although the conditions in Eq. (2) are not satisfied, the ABC estimates lie along the 45 degree line with 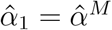; the estimated *x*-effect is nearly unchanged by the addition of the race-modifier. This invariance is *not* satisfied for RGE or STZ. The estimated *x*-effects under RGE or STZ vary considerably between the main-only and race-modified models, with greater discrepancies as the magnitude of the race-modifier effect increases. By comparison, the estimation invariance of ABCs is robust to the magnitude of the race-modifier effect. The mild deviations from the equal-variance condition Eq. (2) are most impactful when *γ* = 1.5, which represents the unusual setting in which the interaction effect is much larger than the main effect. Even in this challenging case, the ABC estimates remain nearly invariant between the main-only and race-modified models, especially when compared to the RGE and STZ counterparts.

### Contrast coding

For OLS estimation, identifiability constraints may be imposed using *contrasts*. In this approach, the linear model is fit under any minimally sufficient identification (RGE, STZ, ABCs, etc.) and the categorical variable coefficients are post-processed using linear contrast matrices. Examples include dummy coding (akin to RGE), effects coding (akin to STZ), weighted effects coding (WEC; akin to ABCs), and Helmert coding (for ordered categories). However, contrasts are typically reserved for main-only models and are difficult to combine with regularized regression and variable selection. Further, these previous approaches do not consider or resolve the inequities of reporting or estimating race-specific effects. In particular, WEC has been advocated only in cases when “a categorical variable has categories of different sizes, and if these differences are considered relevant” (57) or “certain types of unbalanced data that are missing not at random” (58), with regression output that suffers from the same presentation bias that afflicts RGE (59). We do not agree with such restrictions for ABCs, and instead argue that this approach offers an equitable and interpretable parametrization with unique and appealing statistical properties, including both estimation invariance and regularized regression. These estimation invariance results and regularized regression analyses are notably absent from previous contrast coding approaches.

## Data Availability

The North Carolina dataset cannot be released due to privacy protections. However, access to the data can occur through establishing affiliation with the Children's Environmental Health Initiative (contact).

https://www.cehidatahub.org

## Data Availability

The North Carolina dataset cannot be released due to privacy protections. However, access to the data can occur through establishing affiliation with the Children’s Environmental Health Initiative (contact). Additional data documentation is available at https://www.cehidatahub.org.

## Code Availability

The proposed methods are implemented in the statistical software package lmabc in R. This package, along with detailed documentation and examples, is available on GitHub.

## ACKNOWLEDGMENTS

We thank Amy Willis for feedback that greatly improved this work. We also thank Virginia Baskin, Caleb Fikes, Prayag Gordy, and Jai Uparkar for helpful discussions and their contributions to software development. Research was sponsored by the National Institute of Environmental Health Sciences (R01ES028819) and the National Science Foundation (SES-2214726). The content is solely the responsibility of the author(s) and does not necessarily represent the official views of the NIH or the U.S. government. The findings and conclusions in this publication are those of the author(s) and do not necessarily represent the views of the North Carolina Department of Health and Human Services, Division of Public Health.

## A. Supporting Information

**Table A.1.**
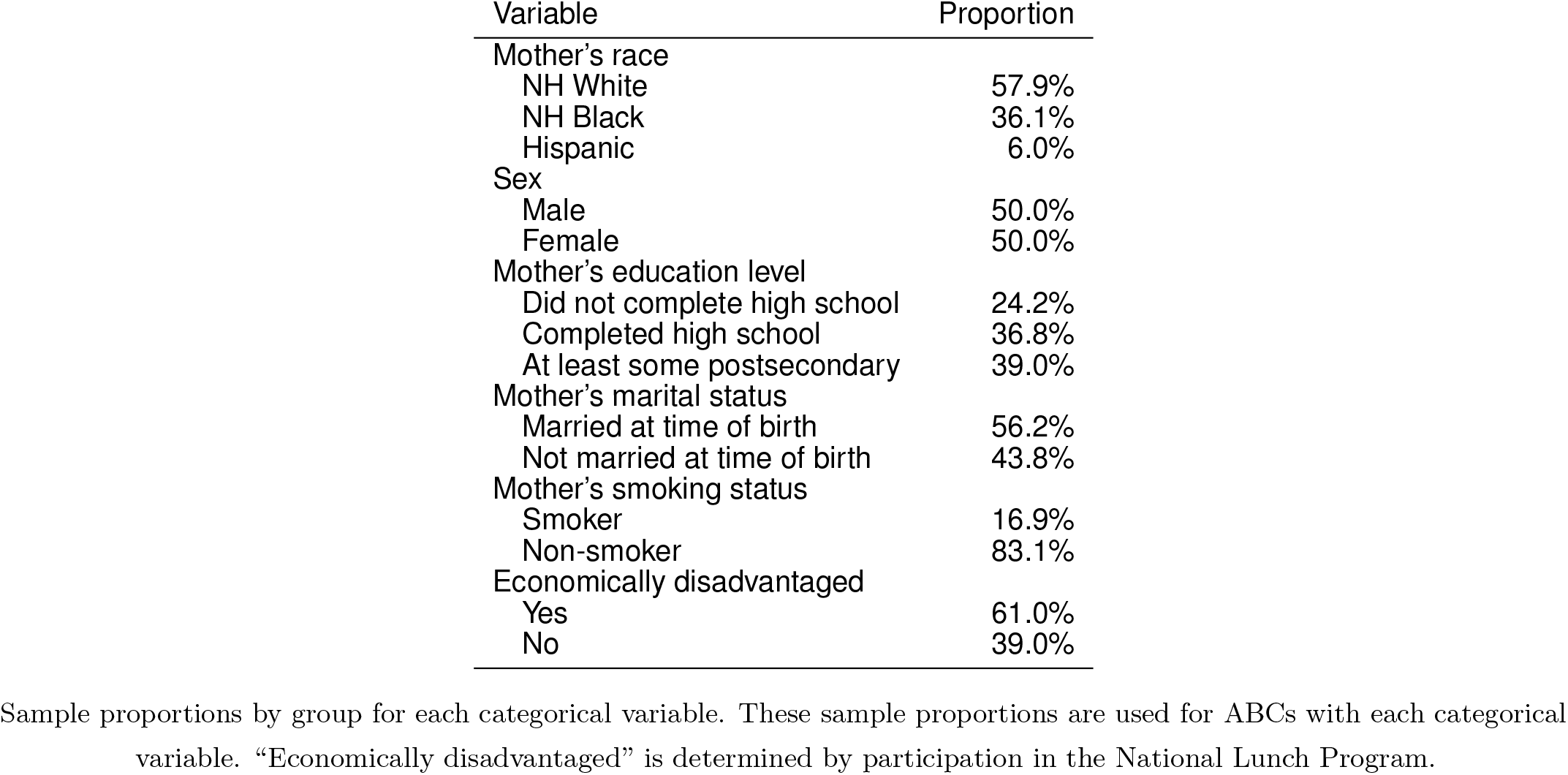
Characteristics of the North Carolina data (*n* = 27, 638).

**Table A.2.** The (scaled) sample standard deviations 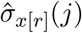 by race *r* for each covariate *j* = 1, …, *p*.

| Variable $j$ | $\hat{\sigma}_{x[\text{NHw}]}(j)$ | $\hat{\sigma}_{x[\text{NHB}]}(j)$ | $\hat{\sigma}_{x[\text{Hisp}]}(j)$ |
| --- | --- | --- | --- |
| Blood lead level | 0.949 | 1.043 | 0.976 |
| PM<sub>2.5</sub> exposure | 0.999 | 1.004 | 0.924 |
| Racial isolation (RI) | 0.688 | 1.063 | 0.936 |
| Mother's age | 0.998 | 0.974 | 0.886 |
| Birthweight percentile for gestational age | 0.998 | 0.955 | 0.984 |

**Fig. A.1.**
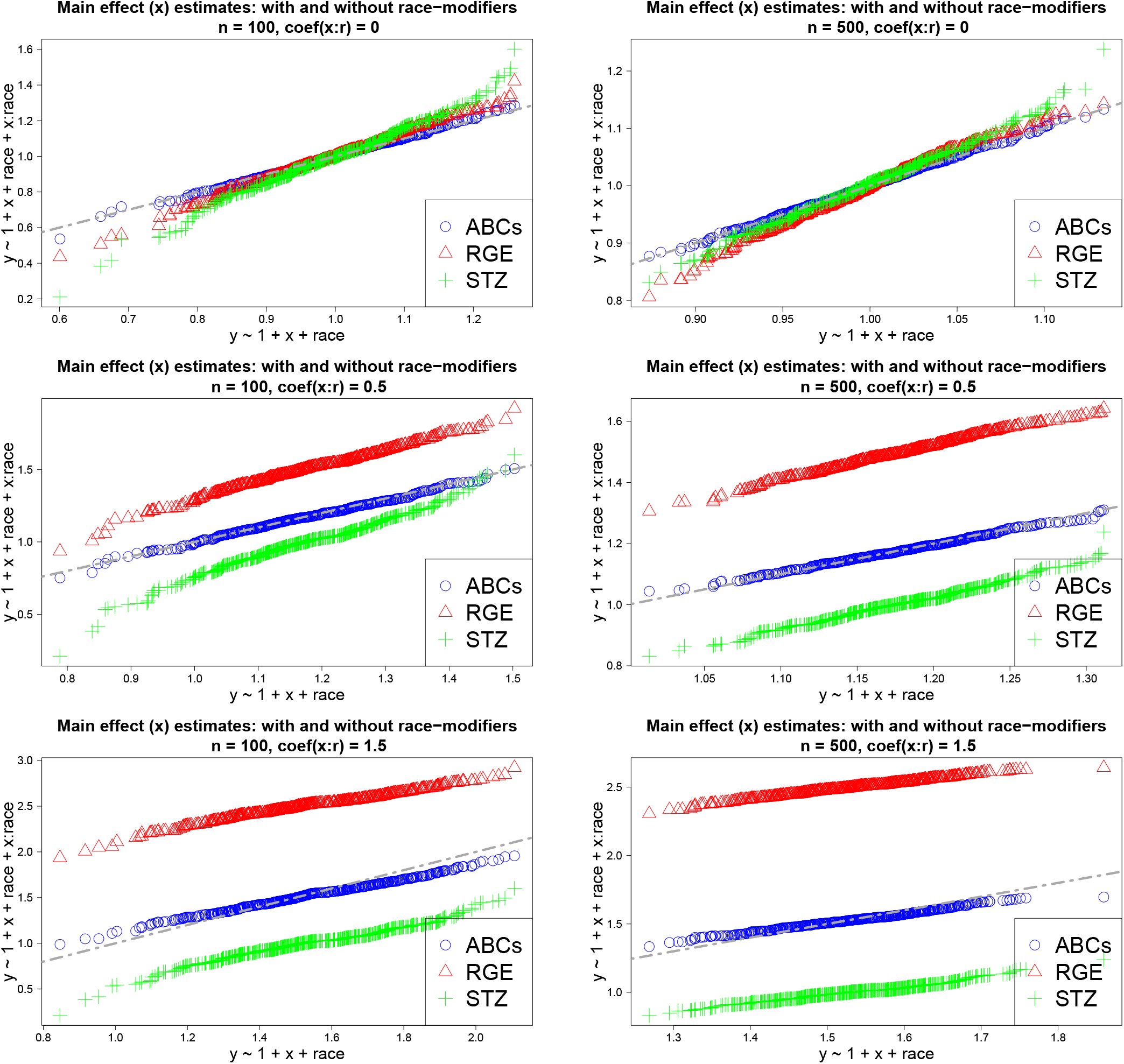
Estimated *x*-effects 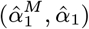 under different categorical encodings across 500 simulated datasets for *n* = 100 (left) and *n* = 500 (right) and varying race-modifier effects *γ* ∈ {0, 0.5, 1.5} (top to bottom). Uniquely, ABCs produce nearly identical *x*-effect estimates with and without the race-modifier (45^°^ line), which preserves the interpretations from the simpler (main-only) model.

**Fig. A.2.**
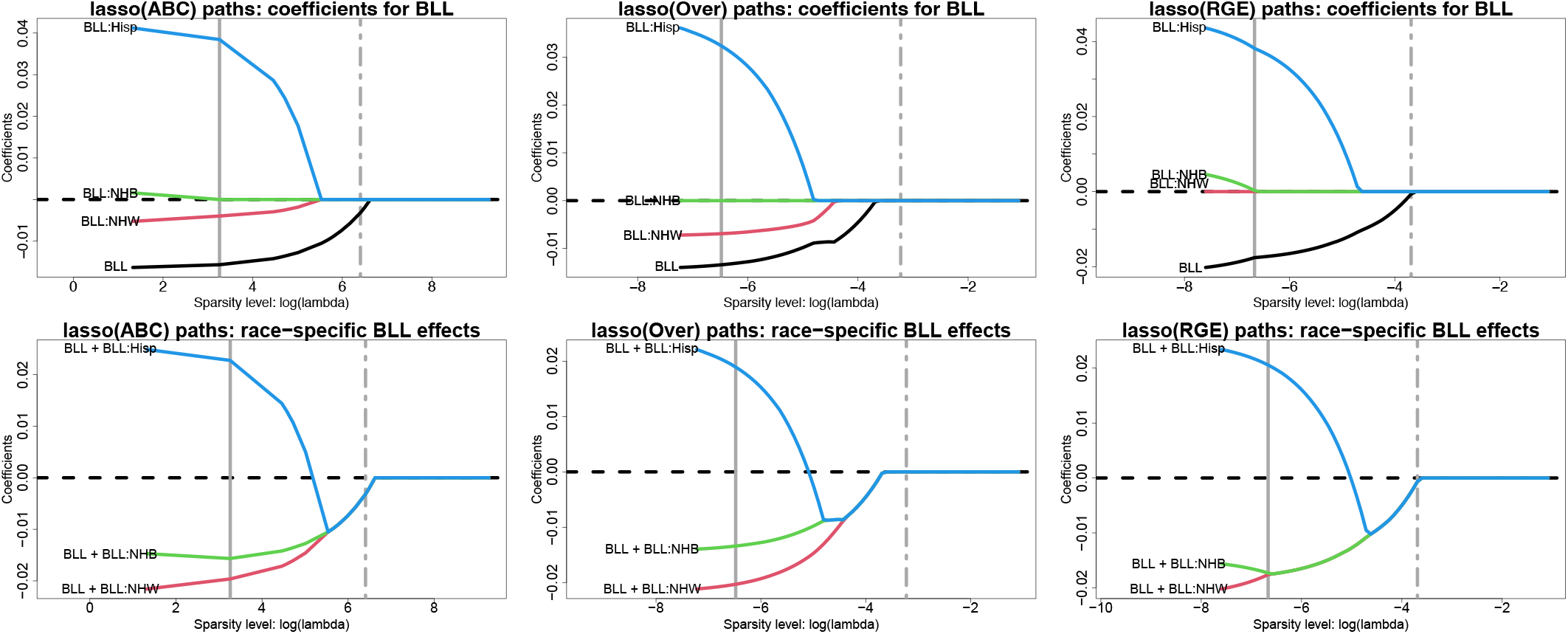
Estimated lasso paths for blood lead level (BLL) across varying sparsity levels (log *λ*) for the model coefficients 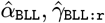 (top) and the race-specific slopes 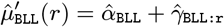 (bottom) under ABCs (left), overparametrized estimation (center), and RGE (right); vertical lines identify *λ* for the minimum CV error (solid) and one-standard-error rule (dot-dashed). The outcome is 4th end-of-grade reading score and the covariates include all variables in Table 2. Small *λ* approximately corresponds to OLS, while increasing *λ* yields sparsity. Under RGE, the race-specific effects are pulled toward the NH White estimate (bottom right). For overparametrized estimation, the paths are similar to the ABC versions (center and left), but estimate 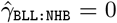 for all *λ* and thus implicitly selects NH Black as the reference group. This explains the differences from RGE, which uses a NH White reference group 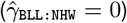. Under ABCs, the race-specific effects are pulled toward a global BLL effect (bottom left), which is is nonzero and detrimental for 4th end-of-grade reading scores.

**Fig. A.3.**
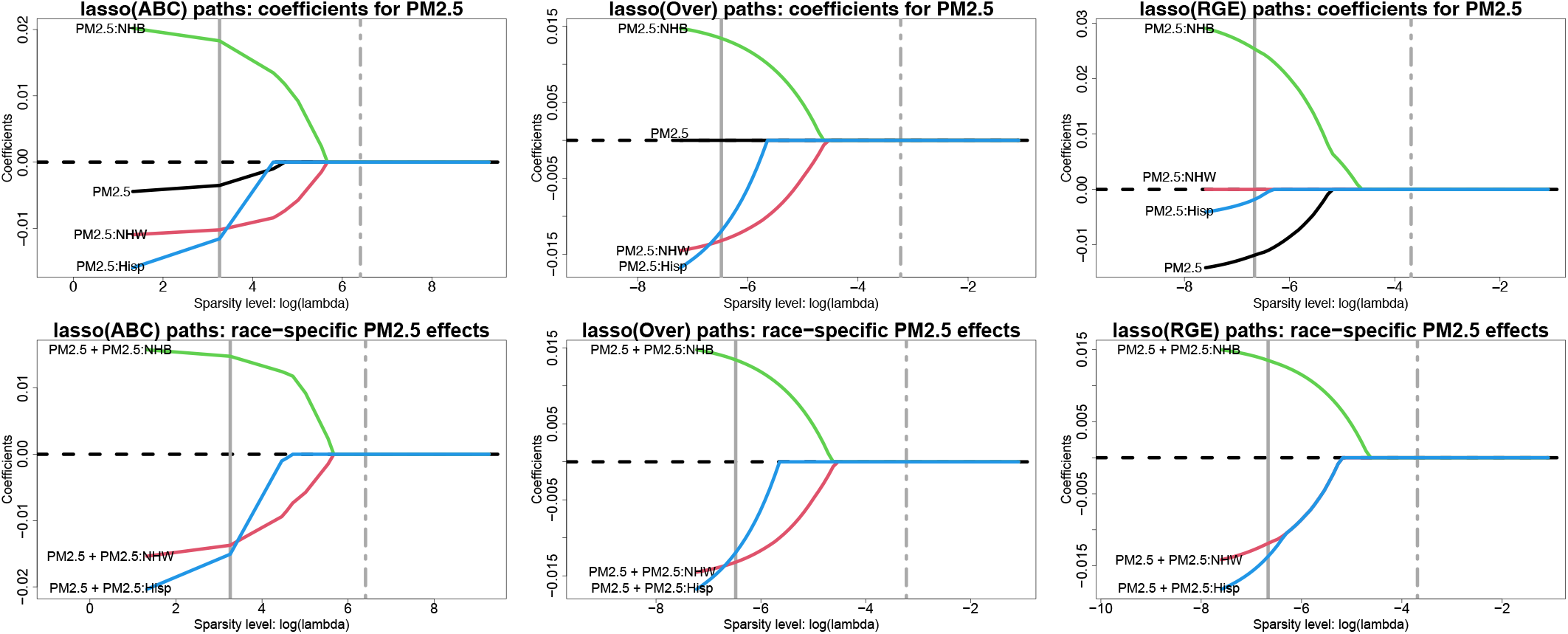
Estimated lasso paths for PM_2.5_ exposure (PM_2.5_) across varying sparsity levels (log *λ*) for the model coefficients 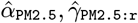 (top) and the race-specific slopes 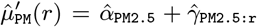 (bottom) under ABCs (left) overparametrized estimation (center), and RGE (right); vertical lines identify *λ* for the minimum CV error (solid) and one-standard-error rule (dot-dashed). The outcome is 4^th^ end-of-grade reading score and the covariates include all variables in Table 2. Small *λ* approximately corresponds to OLS, while increasing *λ* yields sparsity. The ABC paths confirm the OLS output: the global PM_2.5_ effect is pulled toward zero in advance of the race-specific deviations (top left), so the race-specific slopes merge at a global estimate of zero (bottom left). The RGE estimates demonstrate the shrinkage of race-specific effects toward the NH White estimate. The overparameterized paths for the race-specific effects resemble those for ABCs (bottom center and bottom left), but the overparametrized version sets the main effect to zero, 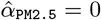 and thus results in different coefficients compared to either ABCs or RGE (top).

**Fig. A.4.**
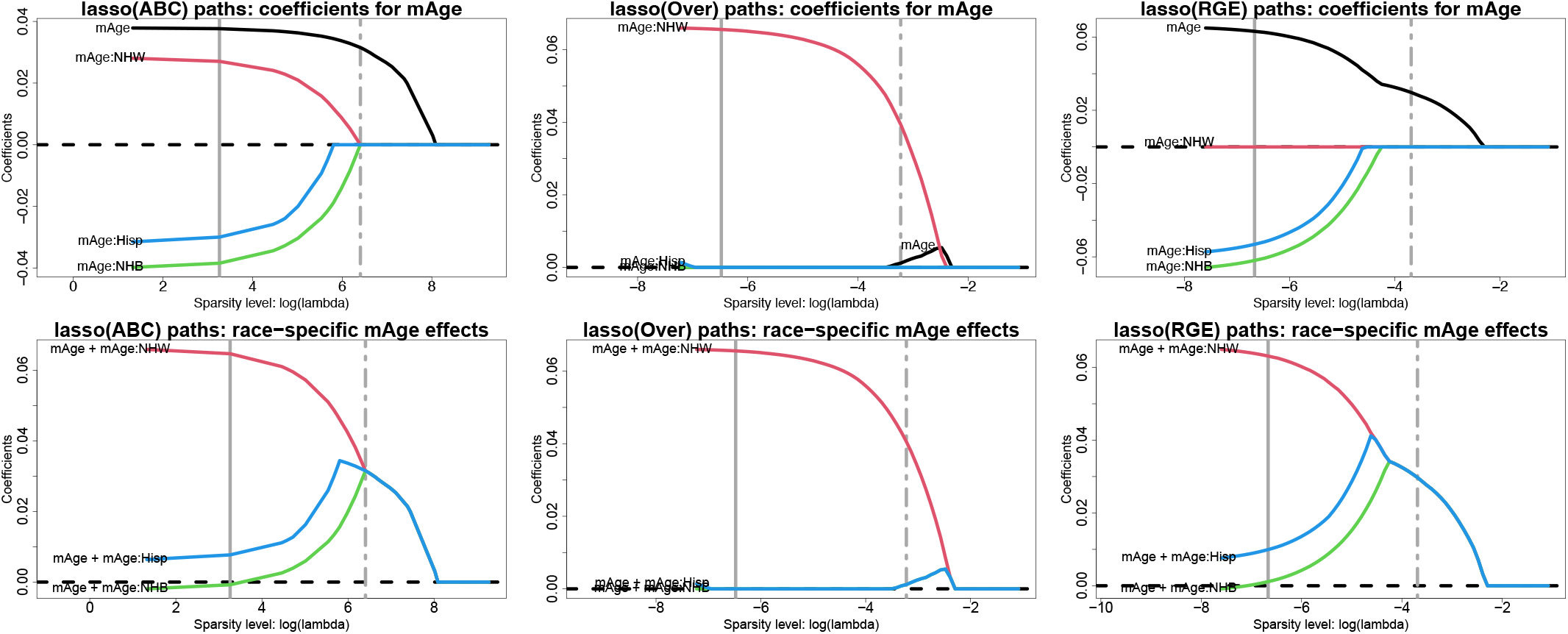
Estimated lasso paths for mother’s age (mAge) across varying sparsity levels (log *λ*) for the model coefficients 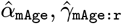 (top) and the race-specific slopes 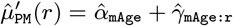 (bottom) under ABCs (left) overparametrized estimation (center), and RGE (right); vertical lines identify *λ* for the minimum CV error (solid) and one-standard-error rule (dot-dashed). The outcome is 4th end-of-grade reading score and the covariates include all variables in Table 2. Small *λ* approximately corresponds to OLS, while increasing *λ* yields sparsity. The racial bias of RGE is clear (bottom right): the race-specific effects are each pulled toward the NH White estimate. By comparison, under ABCs, the race-specific effects are pulled toward a global mAge effect (bottom left), which is nonzero and positive for 4th end-of-grade reading scores. The overparametrized estimation cannot determine a reference group, and exhibits erratic behavior that does not resemble either alternative.

**Fig. A.5.**
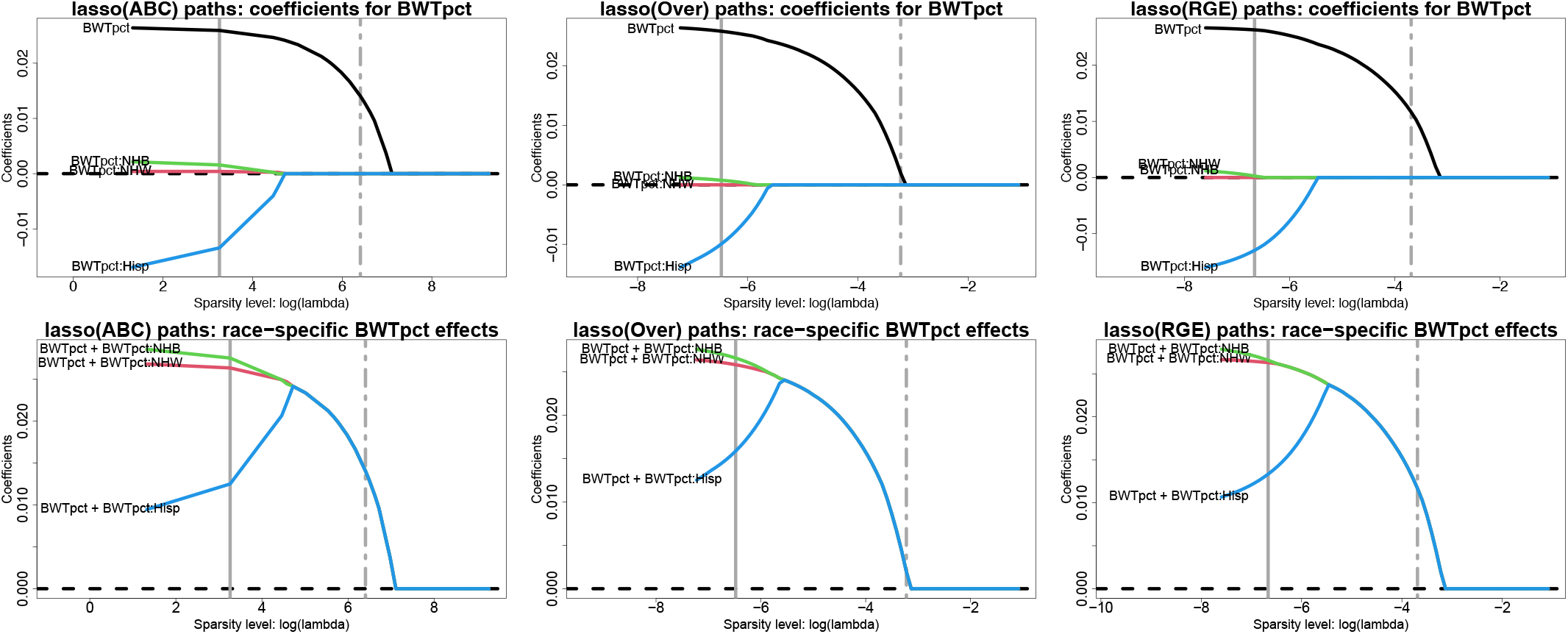
Estimated lasso paths for birthweight percentile for gestational age (BWTpct) across varying sparsity levels (log *λ*) for the model coefficients 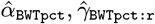 (top) and the race-specific slopes 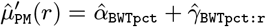 (bottom) under ABCs (left) overparametrized estimation (center), and RGE (right); vertical lines identify *λ* for the minimum CV error (solid) and one-standard-error rule (dot-dashed). The outcome is 4th end-of-grade reading score and the covariates include all variables in Table 2. Small *λ* approximately corresponds to OLS, while increasing *λ* yields sparsity. Under ABCs, the race-specific effects are all positive, and merge at a positive global effect of BWTpct. Because the race-specific deviations for NH White individuals are near zero (top left), the RGE paths—which fix these coefficients at zero by design—are very similar to the ABC paths. This effect is similar for overparametrized estimation.

**Fig. A.6.**
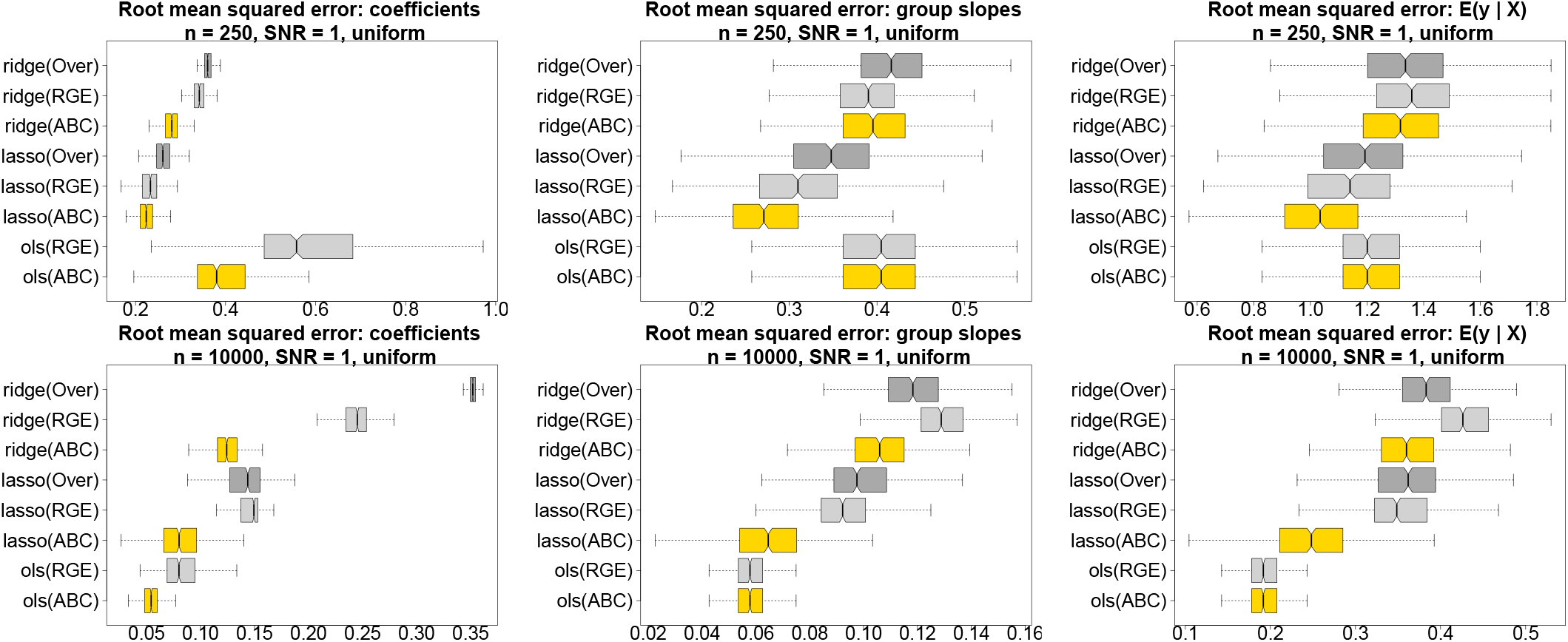
Estimation and prediction accuracy for the regression coefficients (left), the race-specific slopes (center), and the fitted values (right) for *n* = 250 (top) and *n* = 10,000 (bottom) across 500 simulated datasets; nonoverlapping notches indicate significant differences between medians. Data are generated from a Gaussian main-only model with *p* = 10 covariates and a categorical variable with uniform proportions ***π*** = (0.25, 0.25, 0.25, 0.25)^⊤^; both RGE and ABCs are satisfied in the true data-generating process. All fitted models use the race-modified model Eq. (1). ABCs (gold) outperform both RGE (light gray) and `Over` (dark gray) within each estimation method (ridge, lasso, OLS). By definition, the OLS race-specific slopes and fitted values are invariant to the constraints (ABCs or RGE), and `Over` cannot be computed for OLS.

